# Externally Validated Machine Learning Algorithm Accurately Predicts Medial Tibial Stress Syndrome in Military Trainees; A Multi-Cohort Study

**DOI:** 10.1101/2023.01.19.23284808

**Authors:** Angus Shaw, Phillip Newman, Jeremy Witchalls, Tristan Hedger

## Abstract

**Objectives:** Medial Tibial Stress Syndrome (MTSS) is a common musculoskeletal injury, both in sports and the military. There is no reliable treatment and reoccurrence rates are high. Prevention of MTSS is critical to reducing operational burden. Therefore, this study aimed to build a decision-making model to predict the individual risk of MTSS within officer cadets and test the external validity of the model on a separate military population.

**Design:** Prospective cohort study.

**Methods:** This study collected a suite of key variables previously established for predicting MTSS. Data was obtained from 107 cadets (34 females and 73 males). A follow-up survey was conducted at 3-months to determine MTSS diagnoses. Six ensemble learning algorithms were deployed and trained 5 times on random stratified samples of 75% of the dataset. The resultant algorithms were tested on the remaining 25% of the dataset and the models were compared for accuracy. The most accurate new algorithm was tested on an unrelated data sample of 123 Australian Navy recruits to establish external validity of the model.

**Results:** Random Forest modelling was the most accurate in identifying a diagnosis of MTSS; (AUC = 98%). When the model was tested on an external dataset, it performed with an accuracy of 94% (F1= 0.88).

**Conclusion:** This model is highly accurate in predicting those who will develop MTSS. The model provides important preventive capacity which should be trialled as a risk management intervention.

## Introduction

Medial Tibial Stress Syndrome (MTSS) is a common cause of exercise-induced leg pain.^1^ Yates and White define MTSS as “pain along the posteromedial border of the tibia that occurs due to exercise, excluding pain from ischaemic origin or signs of stress fracture.”^2^ Pain is typically spread over a minimum of 5cm, and is recognisable upon palpation of this posteromedial tibial border.^2^ Symptoms are also described as a dull ache following exercise, lasting for hours or days.^2^ In severe cases, pain may be provoked at rest and during activities of daily living.^1, 2^

MTSS is frequently seen in active individuals, including runners, jumping athletes and military personnel.^3^ A 2012 systematic review, which included 3,500 runners identified an incidence of 13.6%-20% over a 12-month period.^4^ This review highlighted MTSS as the most common running-related musculoskeletal injury, ahead of Achilles tendinopathy, plantar fasciopathy and patellofemoral pain syndrome.^4^ MTSS presents a significant medical burden among military populations, with up to 35% incidence reported across a 10-week period.^2^ Prospective data from 6608 British Army recruits found that MTSS is associated with the longest rehabilitation time, accounting for 19.8% of total recovery days.^5^ Australian Defence Force Academy (ADFA), a tri-service officer cadet training facility, injury surveillance data from 2008 showed a mean of 57.5 days of incapacity per individual due to MTSS, which equates to AUD$6,820 of wage costs for working days lost.^6, 7^ In running populations, some runners may even take up to 300 days to recover sufficiently enough to complete an 18-minute run.^8^

Considerable research has investigated risk factors for MTSS.^3, 9, 10^ Many risk factors have been proposed, including leg length, ankle ROM, and smoking history.^9^ Garnock et al. (2018) utilised a suite of risk factors from 2 independent systematic reviews with meta-analyses^9,10^(gender, MTSS history, years of running experience, prior orthotic use, body-mass index (BMI), navicular drop (ND), ankle plantarflexion (PF) and hip external rotation (ER) range of motion (ROM) to produce a statistically significant model, Concordance-Statistic (AUC) = 0.81, for predicting MTSS development.^7^ Their study targeted a predominately male military population of Navy recruits, and thus, the generalisability of the predictive model is still unknown. Understanding the risk factors for MTSS is key, particularly in the absence of a definitive treatment.^1, 10^ Risk factor identification has the potential to be the foundation for designing individualised injury prevention programs, to ultimately help decrease the incidence of MTSS.^10^

The aims of this study were threefold.

1. To train and test a model to predict the individual risk of MTSS in first year ADFA officer cadets.
2. To evaluate the accuracy of the model by external-cross validation on data from a separate military population.
3. To evaluate the accuracy of the model on the two datasets combined.

## Methods

### Study Population

This research used a prospective cohort study design within a sample of volunteer first year ADFA cadets undergoing three-months of initial military training. The study design replicated a previous prospective study at the Australian Navy Recruit School in Victoria, Australia.^7^ The results of this Navy study served as the cross-validation dataset.

The Officer Training College (OTC) at ADFA was selected as the primary site for investigation as it is a tri-service (Army, Navy, and Air Force) military population. Ethics approval was obtained from The Departments of Defence and Veterans’ Affairs (DDVA) Human Research Ethics Committee (HREC) (167-19), and the University of Canberra HREC (20193336).

Participants were recruited face-to-face during induction day in January 2020. Included in the study were trainees, aged 18 years or above, and giving voluntary consent to participate. Participants were excluded if they were currently experiencing shin pain or being treated for MTSS.

## Data Collection

Once written consent was gained from eligible cadets, screening for MTSS risk factors proceeded and included a 4-minute physical examination and a 5-minute paper-based survey.

The physical examination included navicular drop, BMI, and passive ranges of motion (PROM) for ankle plantarflexion and hip external rotation. BMI was calculated after height and weight measurements using a pair of digital scales and a stadiometer (CPWplus 200M Floor Scales, Seca 206 Height Measure). The primary investigator conducted all ankle plantarflexion (PF) PROM measures, whilst another investigator performed all hip ER measures to eliminate inter-rater bias. Hip ER and ankle PF were measured using a digital goniometer (Halo Digital Goniometer, Halo Medical Devices, Sydney, Australia). Digital goniometry has previously been shown to yield high inter-rater reliability with inter-class correlation (ICC) estimates of ICC = 0.99 in Hancock et al. (2018)^11^ and ICC = 0.89 to 0.98 in Correll et al. (2018)^12^.

The measurements of hip ER PROM followed the procedure by Garnock et al.^7^ The measurement of ankle PF was performed with the subject in long sitting. The goniometer stationary axis was aligned with the lateral malleolus, and the fibula and base of the fifth metatarsal served as the moving axis landmark.^13^

Navicular drop was calculated as the difference in height of the navicular tuberosity between relaxed stance and single leg weight bearing.^7^ This process yields moderate intra-rater reliability (ICC = 0.61-0.79)^14^ and a high inter-rater reliability (ICC = 0.84).^15^

A paper-based survey included questions about previous history of MTSS, prior orthotic use and, years of running experience. The additional variables of estimated average number of runs per week and distance per run in the last 6-months were included as they had not been investigated in previous research.^7^ The survey was completed by participants within two-weeks of the initial physical examination.

At three-month follow-up, a paper-based survey captured the incidence and severity of any MTSS cases. Survey items were taken from the MTSS score, including questions on shin pain after exercise (including marching), while walking and at rest.^16^ A physical examination by a Physiotherapist was conducted to confirm any symptoms of MTSS. This examination followed a three-step process (pain history, location of pain and shin palpation).^2^ A systematic procedure was then followed by the Physiotherapist to exclude other lower leg injury syndromes that may masquerade as MTSS.^17^

Strict ethics limitations meant that the researchers were unable to control whether a cadet reported to have their MTSS symptoms confirmed by a Physiotherapist. To account for this, a diagnosis of MTSS was based on one of the two options: (1) either a survey response *and* confirmation by a Physiotherapist with physical examination, (2) or a survey response only, indicating medial border distal tibia pain on self-examination as per Yates and Whites’ definition^2^, associated with walking and/or running.

Statistical analysis examined which combination of the ten MTSS risk factors produced the best predictive model for assessing the risk of future development of MTSS.^7^ Data was analysed using Orange (Data Mining Toolbox in Python developed by Bioinformatics Lab at University of Ljubljana, Slovenia). Variables input to the models were checked for covariance and information gain. Six supervised machine learning methods (Decision Tree, Support Vector Machine, Logistic Regression, K-Nearest Neighbour, Random Forest, Calibrated Random Forest and Naïve Bayes) were used to build predictive models.

Models were trained using five random folds of 75% of the data, and resultant models were cross-validated on the remaining 25% of data in each fold. Each model was then tested for accuracy using the C-Statistic/AUC, classification accuracy (CA), F1, precision, and recall.^18^ The best predictive model identified was tested on the dataset from Garnock et al’ s Navy population.^7^

Using Garnock’ s findings^7^ (AUC = 0.81), power calculations were performed using MedCalc (MedCalc Software Ltd, Belgium). With an estimated MTSS prevalence of 30% MTSS, this study needed 216 participants, with 63 MTSS cases for adequate power.

## Results

During this study, COVID-19 physical distancing restrictions were enforced by the Australian Government. Email correspondence with DHC Physiotherapists notified the research team that cadets were no longer participating in structured physical training sessions. Participants in the study were therefore limited to their ‘ own’ training, which included self-directed running and body weight exercises. Nearby bushfires on the day of initial physical testing also limited availability of volunteers.

Of the 127 cadets that underwent physical screening for MTSS risk factors, a total of 107 (84%) cadets were available to complete the initial MTSS risk-factor survey. Twenty participants were lost to follow-up due to unavailability at the time of survey completion on base. This left 107 cadets (73 males, 34 females) to be followed prospectively across the three-months of initial military training. At three-month follow-up, 99 cadets (69 males, 30 females, mean age 19.3 (Table 1)) remained for inclusion in statistical analysis, with 8 lost to follow-up (unavailable to complete survey).

**Table 1.**
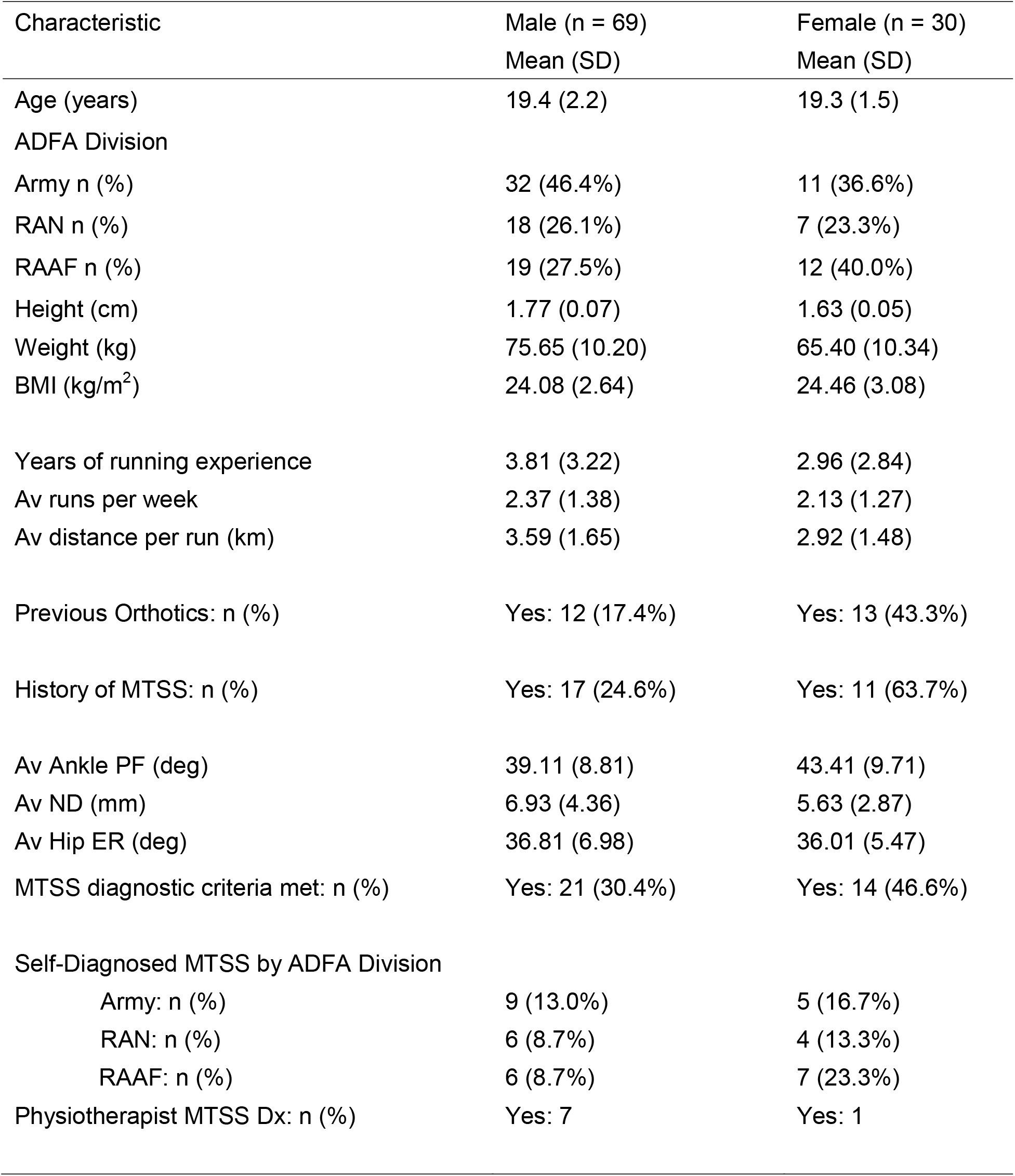
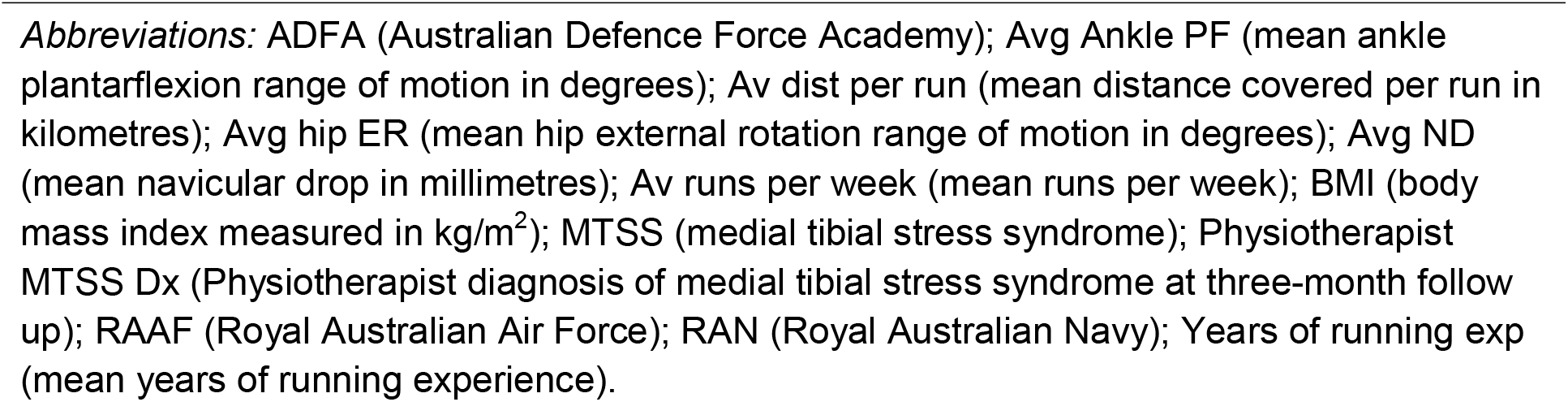
Characteristics of Participants – First Year Australian Defence Force Academy Officer Cadets.

| Characteristic | Male (n = 69)<br>Mean (SD) | Female (n = 30)<br>Mean (SD) |
| --- | --- | --- |
| Age (years) | 19.4 (2.2) | 19.3 (1.5) |
| ADFA Division |  |  |
| Army n (%) | 32 (46.4%) | 11 (36.6%) |
| RAN n (%) | 18 (26.1%) | 7 (23.3%) |
| RAAF n (%) | 19 (27.5%) | 12 (40.0%) |
| Height (cm) | 1.77 (0.07) | 1.63 (0.05) |
| Weight (kg) | 75.65 (10.20) | 65.40 (10.34) |
| BMI (kg/m<sup>2</sup>) | 24.08 (2.64) | 24.46 (3.08) |
| Years of running experience | 3.81 (3.22) | 2.96 (2.84) |
| Av runs per week | 2.37 (1.38) | 2.13 (1.27) |
| Av distance per run (km) | 3.59 (1.65) | 2.92 (1.48) |
| Previous Orthotics: n (%) | Yes: 12 (17.4%) | Yes: 13 (43.3%) |
| History of MTSS: n (%) | Yes: 17 (24.6%) | Yes: 11 (63.7%) |
| Av Ankle PF (deg) | 39.11 (8.81) | 43.41 (9.71) |
| Av ND (mm) | 6.93 (4.36) | 5.63 (2.87) |
| Av Hip ER (deg) | 36.81 (6.98) | 36.01 (5.47) |
| MTSS diagnostic criteria met: n (%) | Yes: 21 (30.4%) | Yes: 14 (46.6%) |
| Self-Diagnosed MTSS by ADFA Division |  |  |
| Army: n (%) | 9 (13.0%) | 5 (16.7%) |
| RAN: n (%) | 6 (8.7%) | 4 (13.3%) |
| RAAF: n (%) | 6 (8.7%) | 7 (23.3%) |
| Physiotherapist MTSS Dx: n (%) | Yes: 7 | Yes: 1 |

*Abbreviations:* ADFA (Australian Defence Force Academy); Avg Ankle PF (mean ankle plantarflexion range of motion in degrees); Av dist per run (mean distance covered per run in kilometres); Avg hip ER (mean hip external rotation range of motion in degrees); Avg ND (mean navicular drop in millimetres); Av runs per week (mean runs per week); BMI (body mass index measured in kg/m<sup>2</sup>); MTSS (medial tibial stress syndrome); Physiotherapist MTSS Dx (Physiotherapist diagnosis of medial tibial stress syndrome at three-month follow up); RAAF (Royal Australian Air Force); RAN (Royal Australian Navy); Years of running exp (mean years of running experience).

During the three-month training period, 35 cadets (35.5%) met diagnostic criteria for MTSS, 21 (30.4%) males and 14 females (46.6%). Calibrated Random Forest modelling (CRFM) yielded the highest AUC, CA and F1 in predicting a diagnosis of MTSS (Table 2). Random forest models were built upon a set of ten decision trees, with each tree developed from a bootstrap sample from 75% of the training dataset. The final predictive model was based on the ‘ majority vote’ from the individually developed trees within the forest.

**Table 2.** Accuracy of Calibrated Random Forest Models (CRFM).

| Model | AUC | CA | F1 | Precision | Recall |
| --- | --- | --- | --- | --- | --- |
| Calibrated RFM trained and tested on ADFA Dataset | 0.98 | 0.96 | 0.96 | 0.96 | 0.96 |
| Calibrated RFM tested on Navy Dataset | 0.95 | 0.94 | 0.88 | 0.89 | 0.86 |
| Calibrated RFM tested on combined Dataset | 0.92 | 0.91 | 0.85 | 0.87 | 0.83 |

**Abbreviations:** ADFA; Australian defence force academy, AUC; Area under curve or C-Statistic CA; Classification accuracy.

Cross validation and testing five times on the remaining 25% of the dataset revealed an AUC of 0.98, CA; 0.96, and F1; 0.96 on average. Testing the CRFM on an unrelated Navy dataset^7^ n=123, 95 males, 28 females, and a total of 30 MTSS cases, revealed comparable accuracy in predicting risk for MTSS (AUC; 0.95, CA; 0.94, F1; 0.88). When the CRFM was tested on the combined ADFA and Navy dataset, it maintained good predictive accuracy with AUC; 0.92, CA; 0.91, and F1; 0.85. (Table 2). Confusion matrix analysis showed the model incorrectly identified 8 out of 222 false positives, and 11 out of 222 false negative predictions. Covariance analysis showed a weak to very weak correlation between the variables. Distance per run and average number of runs per week represented the highest correlation within the model (r = 0.289). MTSS history yielded the strongest information gain on model performance (0.08) (See Supplementary Tables 1 and 2 for correlations and ranking of features within the CRFM).

The risk estimations of the CRFM were visualised via a nomogram produced within the Microsoft Power Business Intelligence interface (Microsoft Corporation, Version 3220.30820.19513.0). Individual risk calculations (minimum, maximum and mean) were modelled by adjusting values of the variables (See Figure 2 for example dashboard).

**Figure 1.**
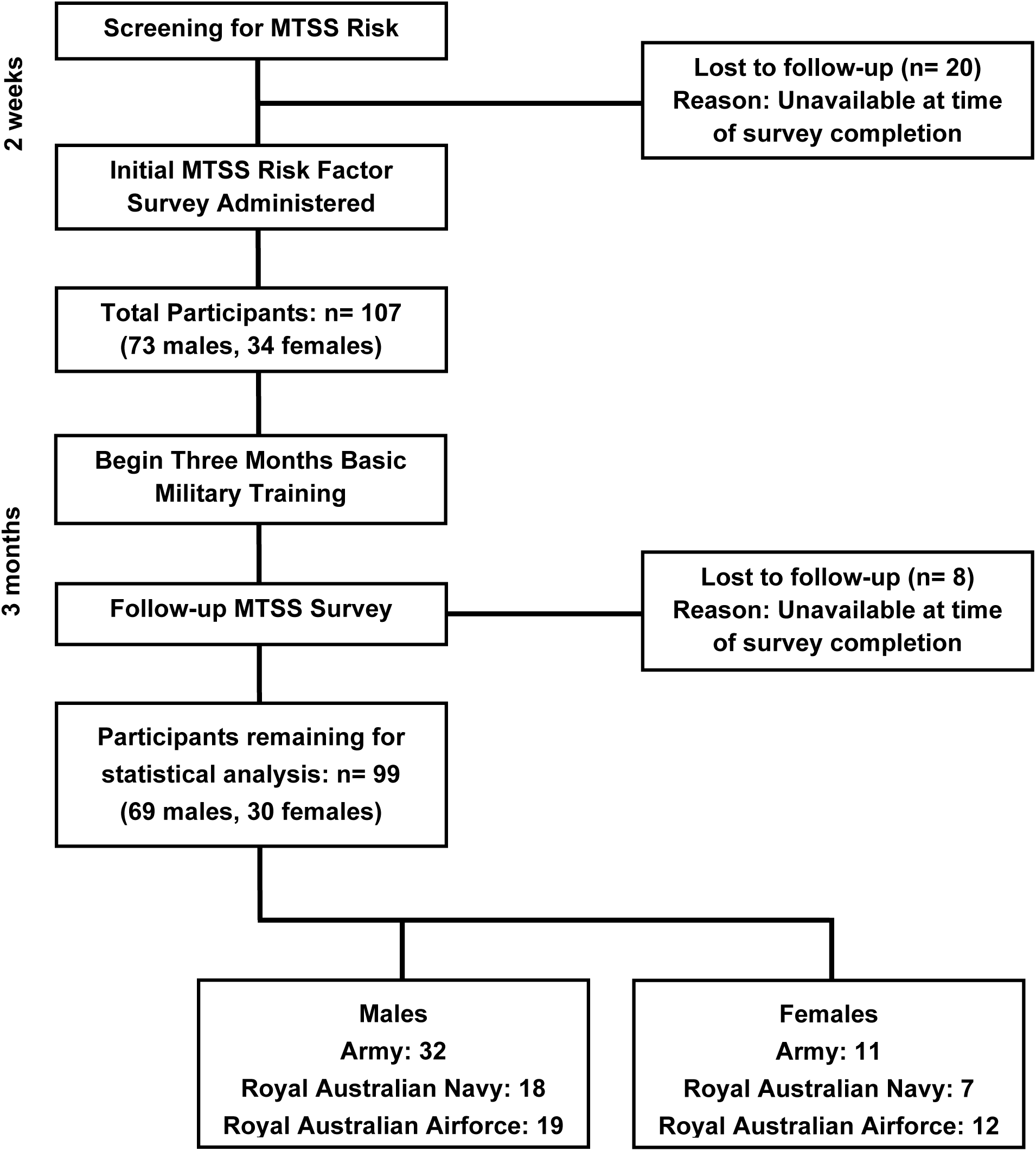
Flowchart of Participants (First Year Australian Defence Force Academy Officer 2 Cadets).

**Figure 2.**
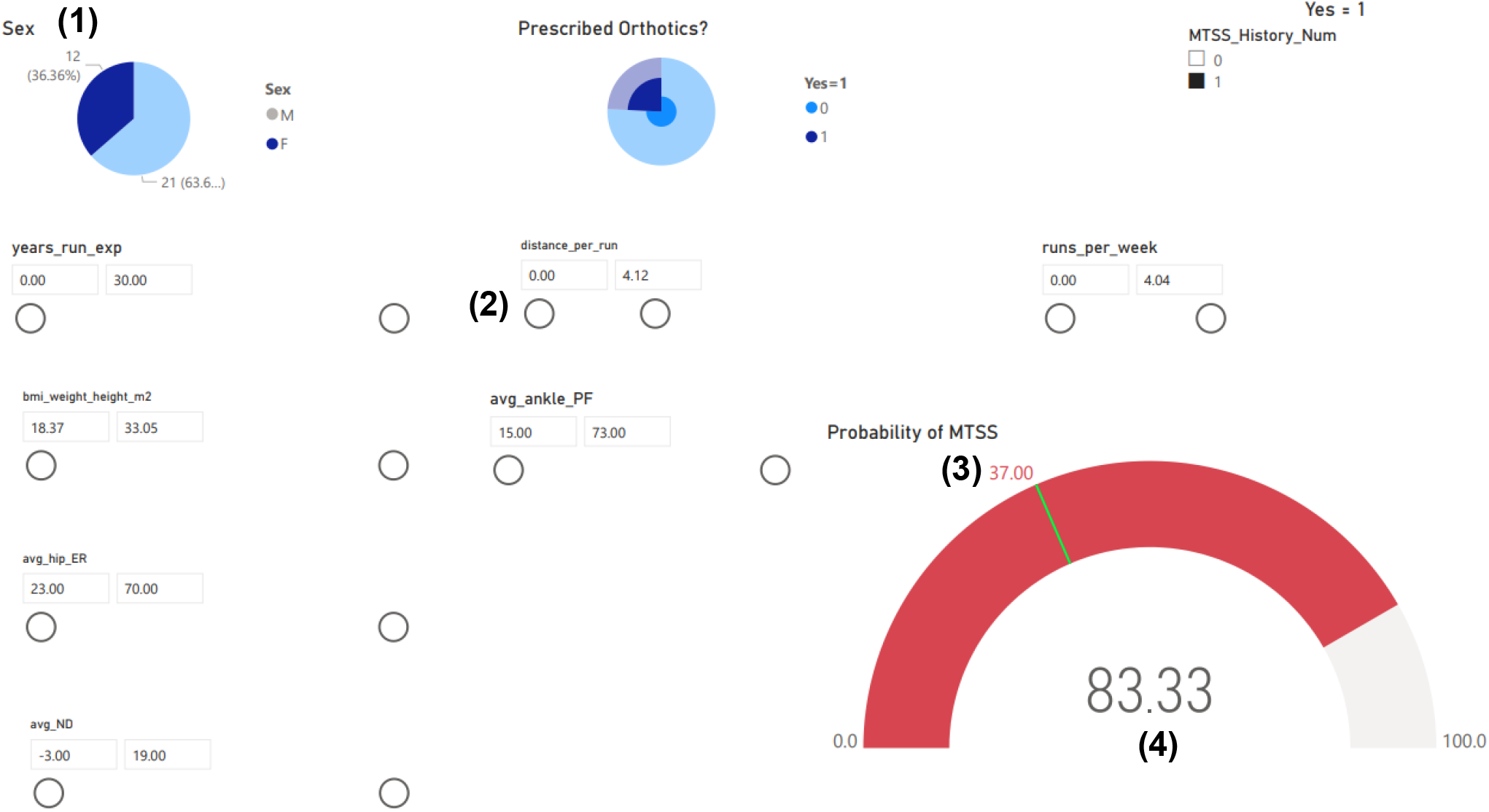
Interactive Nomogram for Calibrated Random Forest Model (CRFM). **Legend;** (1) Allows filtering MTSS risk by males/females (2) Adjustable sliders allowing the dialling up/down of risk factor features (3) Green line represents group probability for MTSS risk (37%) (4) Example scenario; female individual with previous MTSS, who runs four occasions per week at average 4.12 kilometres per run, has an increased MTSS risk of 83.33%.

## Discussion

### Principal findings

This study has developed a predictive model with accuracy of 98% (AUC) when trained and tested on a tri-service dataset. When the model was cross validated against an unrelated dataset from Navy recruits, the model demonstrated a 95% accuracy (AUC). When we applied the model to the two datasets combined, the accuracy remained high, at 92%. The outcomes of this study demonstrate that a model combining a suite of evidence-based risk factors leads to an accurate risk prediction for MTSS. The predictive power of this model appears robust to population change, after validation testing within a separate military.

Having high predictive capacity should facilitate prevention of MTSS. If training, coaching, command staff, or trainees themselves can identify their risk, then risks can be managed. The interactive nomogram (Figure 2) allows individualised risk profiling for MTSS. Probability of MTSS development is calculated based an individual’ s risk factor scores. Once an individual’ s risk of MTSS is calculated, targeting the modifiable risk factors may serve as the strongest preventative measure for this difficult to treat condition. Using this tool, interventions can be modelled and customised to reduce individual risk within their profile of modifiable and unmodifiable characteristics.

This investigation has revealed an MTSS incidence of 35% within a first year ADFA population. This is consistent with previous prospective research in military samples with 24%^7^ and 35% incidence rates^2^. However, the rates of reporting MTSS symptoms to DHC Physiotherapists in this study was low. This isn’ t the first time to have occurred within prospective studies in military personnel. In Yates and White’ s investigation into 124 Naval recruits, only 30% of recruits who developed MTSS followed up with medical treatment.^2^ Furthermore, Almeida et al. investigated gender differences in lower limb musculoskeletal injury reporting rates within a US military population.^19^ A total of 35.8% of injuries went unreported, with MTSS identified as the most common unreported diagnosis.^19^ These findings in combination with the present study may highlight a trend of reluctance to report injuries within the military to avoid being downgraded or medically restricted during their physical training. Giving the cadets the option to have/not to have their MTSS symptoms confirmed by a clinician also likely influenced the rates of reporting. Another reason that may explain underreporting of MTSS symptoms was the COVID-19 physical distancing restrictions. Structured physical training (PT) sessions were ceased. Participants in the study were therefore limited to their own PT, which included self-directed running and body weight exercises. Consequently, there may have been a reduced desire for cadets to present to Physiotherapy to have any underlying MTSS complaints assessed and treated. Out of the 10 features within our model, MTSS history yielded the strongest impact on the performance of the combined CRFM. Having a history of MTSS is a known risk factor for the condition.^7,10^ However, there were very weak correlations between the features, likely signifying the importance for an interaction between all the risk factors combined.

The ADFA first year sample did not reach adequate size for statistical power. Arrivals at data collection day were limited due to bushfire related travel delays. Survey completion and physical training were disrupted due to COVID-19 restrictions. However, the aggregated datasets from ADFA and the Navy trainees did reach adequate sample size with 222 individuals and 65 cases.

### Strength and Limitations

This study has several strengths. The first being the high response rate to the three-month follow-up survey (92.5%). This matches Garnock et al’ s study and represents a highly controlled sample.^12^ This research confirms previous studies^5,7^ investigating MTSS risk in military personnel, with comparable MTSS incidence rates and proportions between sexes. The use of a self-reported survey appears to have increased the compliance with reporting compared to the effort required to visit a physiotherapist. This may have enabled this study to assess the prevalence of a ‘ subclinical’ level of MTSS within this military population. Thus, the chosen approach has value in achieving a broader surveillance of MTSS incidence.

The use of a self-reported MTSS diagnosis as the primary outcome is a key consideration when interpreting the results. The clinical criteria versus self-reported criteria involved the same essential steps; pain history and examination, and self-report has previously shown to reliably match clinician diagnosis of MTSS.^21^ The key difference was Physiotherapist lead versus participant lead palpation for the site of symptoms. The addition of a Physiotherapist examination ensures MTSS symptoms are greater than or equal to 5-centimetres in length and along the posteromedial tibial border.^2^ Physiotherapist examination also assists in ruling out stress fracture or other co-existing injuries.^2, 21^ The prevalence of MTSS within our study was consistent with previous studies where clinical diagnosis was performed, giving further confidence in the results.

#### Future research

This study did not successfully capture week to week training loads, only the self-reported baseline running load. Investigation into risk factor screening combined with training load monitoring to predict future injury is not well established.^22^ Studies have typically used either analysis of risk factors alone, or training loads and their relationship with injury.^7, 23^ It is proposed that those accustomed to increased training loads have reduced injuries compared to those unaccustomed individuals.^22^ In team sport, there is evidence supporting that sharp increases in training load and spikes in acute (7-day) to chronic workload (28-day) ratios (ACWR) are associated with higher injury rates.^24^ Research by Rossi et al.^25^ has shed light on the accuracy of using global positioning systems (GPS) training data in combination with machine learning algorithms to forecast injury risk in team sport athletes. Rossi and colleagues used GPS data to calculate individual ACWR profiles.^25^ However, rather than focusing purely on the ACWR, Gabbett et al.^26^ recommends consideration of known moderators to the workload-injury relationship (e.g., injury history and other factors known to influence the risk of injury). A moderator may either increase or decrease risk of injury at a given training load.^27^ Specific to MTSS, example moderators to the workload-injury relationship may include previous MTSS history and previous years of running experience. Importantly, Wang and colleagues^28^ highlighted several potential limitations with using the ACWR. These authors suggest the ACWR is vulnerable to sparse data bias, time-dependent confounding and recurrent injuries.^28^ An alternative may be the use of causal inference-based strategies, which account for time-dependencies of activity and confounders (e.g. training schedules).^28^ Future research using the ACWR may consider using both time-to-event and multilevel modelling.^29^ Nevertheless, the application of a machine learning methodology which targets risk factor profiling combined with training load monitoring is worth investigating within military settings.^25^

Risk factor profiling for MTSS only contributes to one piece of the puzzle. The next step to managing this difficult condition may be through modifying the modifiable risk factors.^7^ Once an individual has been identified as “at risk”, the design and implementation of injury prevention programs may serve as the best approach to reducing MTSS incidence. Similar to other musculoskeletal conditions, risk prediction for MTSS is often complex and multivariate. In Lahti and colleagues’ work in professional football players, multifactorial and individualised risk reduction programs were prescribed based on the outcomes of risk factor screening.^30^ Such approach is worth investigating within a military sample.

## Conclusion

The outcomes of this study demonstrate that an inexpensive model including a suite of evidence-based risk factors can accurately predict which military trainees will develop MTSS. The model maintains 92% accuracy (AUC) when externally validated in an unrelated military sample. These outcomes enable future research to develop individualised injury prevention programs that address the modifiable MTSS risk factors. The application of predictive modelling methodologies which target risk factor profiling combined with training load monitoring is worth investigating within military settings. Further understanding of how these variables interact and influence MTSS outcomes will help bridge the gap in reducing MTSS incidence.

### What is already known on this topic

Medial Tibial Stress Syndrome (MTSS) is a common musculoskeletal injury in physically active populations, but no reliable treatment(s) exist, and reoccurrence rates are high. Therefore, developing preventative measures are key to reduce injury burden.

### What this study adds

Military institutions, clinicians and instructors are now equipped with a low cost and user-friendly decision-making model, allowing accurate and individual level risk predictions for future MTSS development. The predictive power of the model was proven to be robust to population change, capable of determining MTSS risk within separate military populations.

### How this study might affect research, practice, or policy

Once an individual’ s risk of MTSS is calculated, targeting the modifiable risk factors may serve as the strongest preventative measure for this difficult to treat condition. Using this tool, interventions could be modelled and customised to reduce individual risk within their profile of modifiable and unmodifiable characteristics.

## Data Availability

All data produced in the present study are available upon reasonable request to the authors

## Acknowledgements

We would like the acknowledge the Commanding Officer, all staff, and cadets of the Officer Training College at the Australian Defence Force Academy for their willingness to participate and for their support throughout the study. In particular, the assistance provided by Physiotherapy department at the Duntroon Health Centre during data collection was very much appreciated.

## Contributors

PN and JW contributed to the original study conception and design. Methodology: AS, PN and JW, Data collection: AS, PN, JW, and TH, Statistical Analysis: AS, PN and JW, writing original draft: AS, Writing review and editing: AS, PN, JW, and TH, Supervision: PN and JW.

## Participant consent for publication

Gained.

## Ethics Approval

The Australian Defence Human Research Ethics Committee for a low-risk project; Approval Number 167-19. The University of Canberra Human Research Ethics Committee; Approval Number 20193336.

