## Supplementary Table 1 & 2 for "Externally Validated Machine Learning Algorithm Accurately Predicts Medial Tibial Stress Syndrome in Military Trainees; A Multi-Cohort Study"

Supplementary Table 1. Features Ranked by Contribution to Combined Calibrated Random Forest Model (Information Gain).

| Feature | Information Gain |
| --- | --- |
| MTSS history  Years run experience | 0.0785 0.0327 |
| Sex Average ankle plantarflexion | 0.0310 0.0205 |
| Average hip external rotation | 0.0142 |
| Body mass index Average runs per week  Orthotic history  Average navicular drop  Average distance per run | 0.0111 0.0093 0.0039 0.0038 0.0009 |

Abbreviations: MTSS; Medial Tibial Stress Syndrome

Supplementary Table 2. Feature Correlations within Combined Calibrated Random Forest Model (CRFM).

| Feature 1 | Feature 2 | Correlation (r) |
| --- | --- | --- |
| Average distance per run  Average ankle plantarflexion | Average runs per week  Average runs per week | 0.289 -0.152 |
| Average distance per run  Average ankle plantarflexion | Years run experience  Average hip external rotation | 0.151 0.148 |
| Average ankle plantarflexion | Average distance per run | -0.138 |
| Average hip external rotation  Average runs per week  Average navicular drop  Average ankle plantarflexion  Body mass index | Average runs per week  Years run experience Average ankle plantarflexion Years run experience Average runs per week | 0.108 0.103 -0.078 -0.076 0.076 |
| Average hip external rotation | Average distance per run | -0.068 |
| Average navicular drop | Years run experience | -0.06 |
| Average navicular drop  Body mass index | Body mass index Average distance per run | 0.04 0.026 |
| Average navicular drop  Average hip external rotation | Average runs per week Years run experience | 0.017 -0.016 |
| Average hip external rotation | Body mass index | 0.014 |
| Body mass index  Average navicular drop  Average ankle plantarflexion  Average navicular drop | Years run experience Average distance per run  Body mass index  Average hip external rotation | -0.011 0.005 0.003 -0.002 |
